# Development Process of a Clinical Decision Support System for Empiric Antibiotic Therapies in Sepsis Patients

**DOI:** 10.1101/2025.05.28.25328512

**Authors:** Sophie Schmiegel, Hannah Marchi, Philipp Hege, Svenja Elkenkamp, Juliane Düvel, Christoph Düsing, Wolfgang Greiner, Sean Selim Scholz, Dominic Witzke, Johannes J. Tebbe, Michael Wehmeier, Olaf Kaup, Rainer Borgstedt, Sebastian Rehberg, Philipp Cimiano, Christiane Fuchs

**Author notes:** Corresponding author: Christiane Fuchs. both authors contributed equally (Sophie Schmiegel, Hannah Marchi, Christiane Fuchs).

## Abstract

**Background:** The principal treatment against bacterial infections are antibiotic therapies. However, increasing antibiotic resistances pose a major threat to global health care systems by which sepsis patients are particularly affected. Those patients urgently need to be treated with the most effective antibiotic therapy to maximize their chances of survival while simultaneously preventing the development of both individual and global resistances. Consequently, in order to select a proper empiric antibiotic therapy, the treating physicians need to account for many different factors. A clinical decision support system (CDSS) aims to support physicians in deciding on a fast and targeted antibiotic therapy.

**Objective:** The purpose of this work is to explore the extent to which the realization of a CDSS is possible based on the data available to us, and to document our insights gained during the development of a foundational model designed to assist physicians in determining empiric treatment options for sepsis patients. In this regard, we aim to highlight the importance of close interprofessional collaboration between scientists from various disciplines and to analyze the effects of data quality and quantity on the performance of our statistical models.

**Methods:** Empirical scientists regularly conducted interviews with medical practitioners in order to acquire medical knowledge required to develop sound statistical models. We developed and applied two-step cross-sectional as well as time series classification models to carefully preprocessed data of sepsis patients admitted to the intensive care unit of a German hospital.

**Results:** We identified several factors as crucial information for valid decisions on empiric therapy for treating sepsis patients. These include the patients’ core data, especially the infection focus. To prevent further resistances, individual risk factors such as travel history and professional background should be considered. The evaluation of a therapy’s effectiveness is mainly based on the patient’s general condition and blood values such as procalcitonin and interleukin 6. One key factor in the acceptance of CDSS is the explainability of the results produced by the applied methods. Our models come along with mainly moderate but comprehensive predictive ability for all considered empiric antibiotic therapies.

**Conclusion:** This work highlights the importance of interprofessional collaboration between medical experts and model developers, ensuring that data quality and clinical relevance are central to the process. It emphasizes the urgent need for high-quality, comprehensive data to overcome challenges such as data discontinuity and improve model performance, particularly through enhanced digitization in healthcare. This foundational work will facilitate future efforts to develop a CDSS for treating sepsis patients and to translate it to clinical use.

## 1 Introduction

### 1.1 Motivation

Antibiotic therapies represent a milestone within pharmacological and clinical research and are commonly employed in the treatment of bacterial infections ranging from mild cystitis to life-threatening sepsis. Sepsis is a severe and rapidly progressing course of a bacterial infection which needs to be treated within the first three hours after diagnosis to ensure a patient’s survival [1]. The chance of treatment success rises with increased coverage of pathogens; thus, the Surviving Sepsis Campaign Guidelines for Management of Sepsis and Septic Shock 2021 [1] recommend a broad-spectrum antibiotic as empiric (i. e. the initial) therapy. Accordingly, German hospital guidelines recommend the use of a broad-spectrum antibiotic, in particular if the focus of infection is unknown. While broad-spectrum antibiotics are likely to be effective, they come along with a variety of side effects like gastrointestinal complaints and drug fever [2], and harbor the risk of developing antibiotic resistances [3, 4]. Such resistances pose a global threat to healthcare systems [5, 6]. Thus, an important goal is to minimize the delivery of broad-spectrum antibiotics. A direct point of action for this are inappropriate prescriptions, e. g. in case of viral infections [7], and still common self-prescriptions due to low-threshold access to antibiotics [8]. Overall, preventing the development of further resistances requires consistent antibiotic stewardship and support in selecting targeted antibiotic therapies. The latter is particularly difficult in the case of severe disorders such as sepsis, which are the focus of this work. Even for experienced physicians, the diagnosis and treatment of sepsis is a challenging task [9].

### 1.2 Background

#### 1.2.1 Project Background

This work has emerged from the interprofessional project ‘KINBIOTICS’ [10], funded by the German Federal Ministry of Health from 2020 to 2024. An overarching objective of the project was to develop an artificial intelligence (AI)-based CDSS for the individualized prediction of targeted effective antibiotic therapies with low side effects for patients diagnosed with sepsis. Such a CDSS would help clinicians make rapid and targeted treatment decisions and thus save lives and reduce the progressive occurrence of antimicrobial resistances. Additionally, the process of developing a data-driven CDSS has the potential to reveal previously unconsidered patterns in the data, thereby providing substantial benefits to the current prescription process.

Research in the ‘KINBIOTICS’ project was conducted by medical practitioners and empirical scientists from various disciplines including information technology, data science, biotechnology, health sciences and medicine. The scientists were affiliated to five institutions, namely Bielefeld University, University of Siegen, Evangelisches Klinikum Bethel (EvKB), Klinikum Bielefeld and Klinikum Lippe. The project was organized in work packages addressing a range of topics, including the prediction of the pathogen and resistance spectrum by nanopore sequencing [11], the setup of a data warehouse including the creation of preconditions for federated learning [12], the development of a resistance observatory [13], evaluation of a pilot CDSS [14], statistical model building as basis for the CDSS and the support system’s integration into clinical routine [15].

To achieve the project goals, model constructors and medical professionals were in close dialogue with each other, and this turned out to be particularly helpful for the process of data selection, data preprocessing and statistical model fitting. In the following, we focus on this process and the steps we found essential to be taken prior to the development of an AI-based CDSS.

#### 1.2.2 Sepsis

Sepsis is one of the main causes of illness and death all over the world. In 2017, an estimated 48.9 million incident cases of sepsis were recorded worldwide and 11.0 million sepsis-related deaths were reported, representing 19.7 % of all global deaths [16]. Therefore, the World Health Organization (WHO) urged their members with a resolution to include prevention, diagnosis and treatment of sepsis in national health systems and to develop and implement standard and optimal care for diagnosing and managing sepsis in health emergencies [17].

Sepsis has been defined as a life-threatening organ dysfunction caused by a dysregulated host response to infection (Sepsis-3 Definition) [18]. The infection can be caused by most types of microorganisms, bacteria, fungi, viruses and parasites [19]. Organ dysfunction is characterized using the sequential organ failure assessment (SOFA) score [20], including six organ systems (respiratory, coagulation, liver, cardiovascular, central nervous system and renal) with a rise of at least two points following the infection. The most severe state of sepsis is defined as septic shock with a persistent hypotension requiring vasopressor medication and a serum lactate level of more than 2 mmol/L despite adequate volume resuscitation. This status is characterized by circulatory, cellular and metabolic abnormalities leading to an increased mortality of up to 40 % [18].

According to the resolution of the WHO, international scientific organizations started the Surviving Sepsis Campaign in 2002, developing guidelines for the treatment of sepsis and septic shock [1]. Regarding these guidelines, sepsis and septic shock are medical emergencies requiring rapid diagnosis and immediate treatment. Consequently, the following measures should be carried out as quickly as possible: measurement of the lactate level, commission of an antibiotic susceptibility testing, administration of broad-spectrum antibiotics, initiation of appropriate fluid administration and vasopressors if necessary, and re-measurement of the lactate level. In particular, the administration of antibiotics should not be delayed or miss the pathogen. The rationale behind these measures is that the time until effective antimicrobial therapy is a crucial determinant of survival in septic shock [21]: a delay in the initiation of effective antimicrobial therapy is associated with a mean decrease in survival of 7.6 % per hour.

In clinical practice, the microbiological proof of the pathogen causing sepsis or septic shock is most often missing at the time of diagnosis. Therefore, the initial administration of antibiotics is empirical and generally follows the Tarragona strategy first described by [22]. The principles of this therapy involve assessing the patients and their risk factors, to ’hit hard and early’, aiming at an immediate administration of an effective dose of antibiotics while considering the local antimicrobial situation. It also includes accurately identifying the site of infection and understanding the pharmacodynamics and pharmacokinetics of the antibiotics. Furthermore, it is essential to remain focused by continually re-evaluating the effectiveness of administered antibiotics. Finally, the treatment should be de-escalated accordingly after identifying the pathogen.

### 1.3 Focus of this Work

Many disciplines aim to improve antibiotic therapies, including microbiology, pharmacology, chemistry, and clinical research. Research and knowledge transfer can be significantly more effective when they operate across disciplinary boundaries and connect fundamental research with clinical practice. In our work, we specifically focus on the interprofessional work between scientists from various disciplines with empirical and clinical background aiming at a clinical decision support system (CDSS) for antibiotic treatment; that is an information system designed to assist physicians in making empiric treatment decisions for sepsis patients. The particular value of our contribution arises from its close collaboration between disciplines and through the integration of guidelines, constraints and clinical experience into data analysis. The need for such tailored CDSS continues to exist as, despite all technological advancements, AI systems such as general large language models are still incapable of capturing complex patient situations based on the available data to the extent that they can provide reliable recommendations [23].

On our way to develop such a CDSS, we faced many hurdles to overcome, and this article describes our learnings to support projects with similar objectives in this regard. We describe preparatory steps required prior to the construction of statistical models used for CDSS, and model the physicians’ decision for an empiric antibiotic therapy. In this regard, we highlight the relevance of high data quality and a close exchange between medical experts and model constructors.

### 1.4 State of Research

The causes, diagnosis, treatment and consequences of sepsis are highly relevant in various fields of research. In line with the focus of our work, machine learning and deep learning have commonly been applied to analyze data of sepsis patients. In particular, numerous studies cover the early and accurate prediction of the onset of sepsis, for example [24], [25], [26], [27], and [28]. [29] provide a systematic literature review for early prediction of sepsis in intensive care units (ICU). Further, many studies pursue the goal of predicting antimicrobial resistances. The literature review by [30] identified twelve studies that employ machine learning in the context of antibiotic susceptibility pattern prediction (e. g. [31], [32], and [33]). Additionally, [30] selected eight studies on machine learning-assisted antibiotic resistance prediction (e. g. [34], [35] and [36]). [37] developed a computerized decision support system to predict appropriate antibiotic therapies to avoid the use of broad-spectrum antibiotics for treatment of infectious diseases. Besides, they consider benefits of antibiotics (e. g. reduction in the length of a hospital stay) as well as costs (e. g. adverse events). Such CDSS could also be beneficial in case of sepsis patients, in particular, in assisting physicians in selecting a proper empiric antibiotic therapy for sepsis patients.

## 2 Methods

### 2.1 Data Acquisition and Preprocessing

#### 2.1.1 Medical Knowledge Compilation

As motivated before, in this we attributed great importance to interprofessional collaboration to appropriately meet the challenges of adequate treatment of sepsis, a multi-faceted complication of an infection. Consequently, medical practitioners and empirical scientists maintained continuous dialogue to ensure a deep and comprehensive understanding of both the medical processes and the methodological requirements. This knowledge exchange aimed to guarantee that decisions made during data preprocessing and statistical model construction were congruent with the clinical procedure.

On the operational level, the empirical scientists conducted a series of interviews with predefined questions with representatives of the three participating hospitals, including physicians working at the bedside of sepsis patients. This way, non-clinicians were given a comprehensive overview of treatment options and guidelines, and relevant stakeholders were engaged at all stages of the CDSS development process.

In an initial interview series, two aspects were of primary interest: firstly, the nature and use of broad-spectrum antibiotics, along with aspects of combination therapies; secondly, the evaluation process for the effectiveness of administered therapies, and the critical factors that influenced this evaluation. In two further series of interviews, we focused on relevant determinants of successful implementation of a CDSS in clinical routine settings: by assessing the clinicians’ general acceptance of such a system, and by identifying necessary information to be provided in order to make it as trustworthy as possible. In addition to the interviews, we conducted biweekly meetings between representatives from the hospitals and data scientists. Further, all three hospitals provided their internal guidelines for antibiotic administration and dosage depending on diagnosis and present pathogens. This facilitated the data scientists’ understanding of the specific procedures and decision-making processes. Figure 1 illustrates the process of the entire project under special consideration of statistical model development.

**Figure 1:**
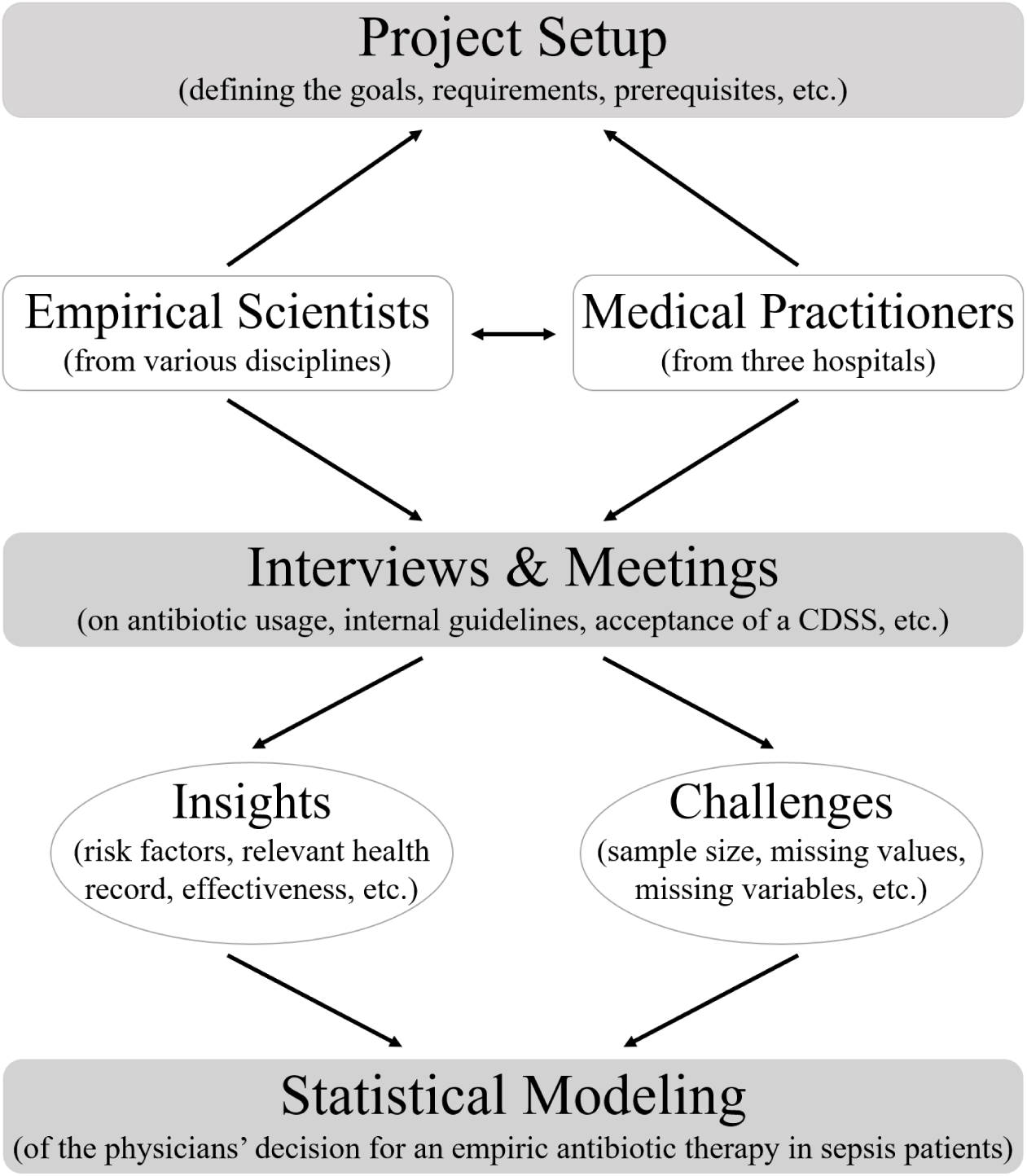
Schematic representation of the process within the ‘KINBIOTICS’ project aiming at modeling the physicians’ decision for an empiric antibiotic therapy in sepsis patients.

#### 2.1.2 Data

We use retrospective health records from patients admitted to the ICU of the German hospital EvKB between 2012 and 2023. All patients in this dataset were diagnosed with sepsis or septic shock. The data was derived from three sources: first, the clinical information system (CIS) included time-constant core data and diagnoses, as well as blood values and vital signs measured over the period of hospitalization. Second, the patient data management system contained administered catecholamines and antibiotics over time. Third, information about microbiology analyses was available from the hospital’s laboratory, representing results of antibiotic susceptibility testing based on patient samples such as blood, urine or smear. However, data of administered catecholamines and microbiological analyses was not included in the analyses as microbiology data, in particular, is not available at the time of diagnosis.

Prior to data analysis, we carefully preprocessed the data. As a first step, we excluded patients who did not receive any antibiotic therapy or who had not yet reached the age of 18 at the time of admission. In collaboration with medical experts, erroneous values in variables such as age, height and weight were either adjusted or removed, and thresholds for blood values and vital signs were included in accordance with medical recommendations. The suspected focus of infection was not documented in the available data. However, given its potential importance for addressing the research question, we derived the suspected infection focus from patients’ ICD-10 codes (International Classification of Diseases, [38]). To this end, a medical expert categorized all sepsis-related ICD-10 codes from the CIS database according to their infection focus. This procedure resulted in many patients having more than one suspected infection focus. Consequently, we split the foci per patient into several variables, taking into account the order of occurrence. For further analysis, however, we exclusively used the first focus and refer to it as the suspected infection focus in the following. A complication occurred by the fact that the time point of diagnosis was only insufficiently documented in the patients’ records. Consequently, we assumed that the time of sepsis diagnosis corresponded to the time of the initial antibiotic administration. Additionally, some patients were represented several times in the dataset. They were either repeatedly admitted to the hospital, or they had multiple infections during one hospital stay. We divided one hospital stay into two separate stays when no antibiotic was administered for at least five days and if there was a change in the infection focus during that time. Henceforth, for the purpose of simplicity, we use the term ’patients’ in place of ’infections’, even in cases where a single patient is represented by multiple infections in the dataset.

The obtained dataset is presented as follows: core data is available for 2,986 patients and contains 26 variables. The age of the patients ranges from 18 to 99, with an average age of 67.56 years; approximately 36 % of patients are female. 1,257 (42 %) of all patients died during the hospital stay. The aforementioned categorization to derive the suspected infection focus ultimately yields the following 16 categories (with the number of occurrences in brackets): lungs (1074), urogenital tract (498), gastrointestinal tract (353), prosthesis inflammation (175), skin/muscle (118), bile (101), brain (82), bones/joints/tendons/intervertebral discs (60), heart (34), vessels (14), liver (11), ear/nose/throat (10), oesophagus (8), oral cavity (3), spleen (1), and eye (1). For all patients, time series of vital signs and blood values (31 variables) are available from a maximum of 88 days before and up to 1,610 days after the date of diagnosis. However, the measurement and documentation of time series variables was carried out with varying quantity and frequency. Table 1 lists all available variables. Diagnoses are documented for 2,945 patients with 3,319 unique ICD-10 codes in total; 24 codes of these connected to sepsis or septic shock. Further, the data contains information about administered antibiotics, including drug names as well as therapy start and end dates, for all patients. A total of 35 distinct antibiotics were administered at least once. An overview of all antibiotics and their frequencies is provided in Figure 2.

**Figure 2:**
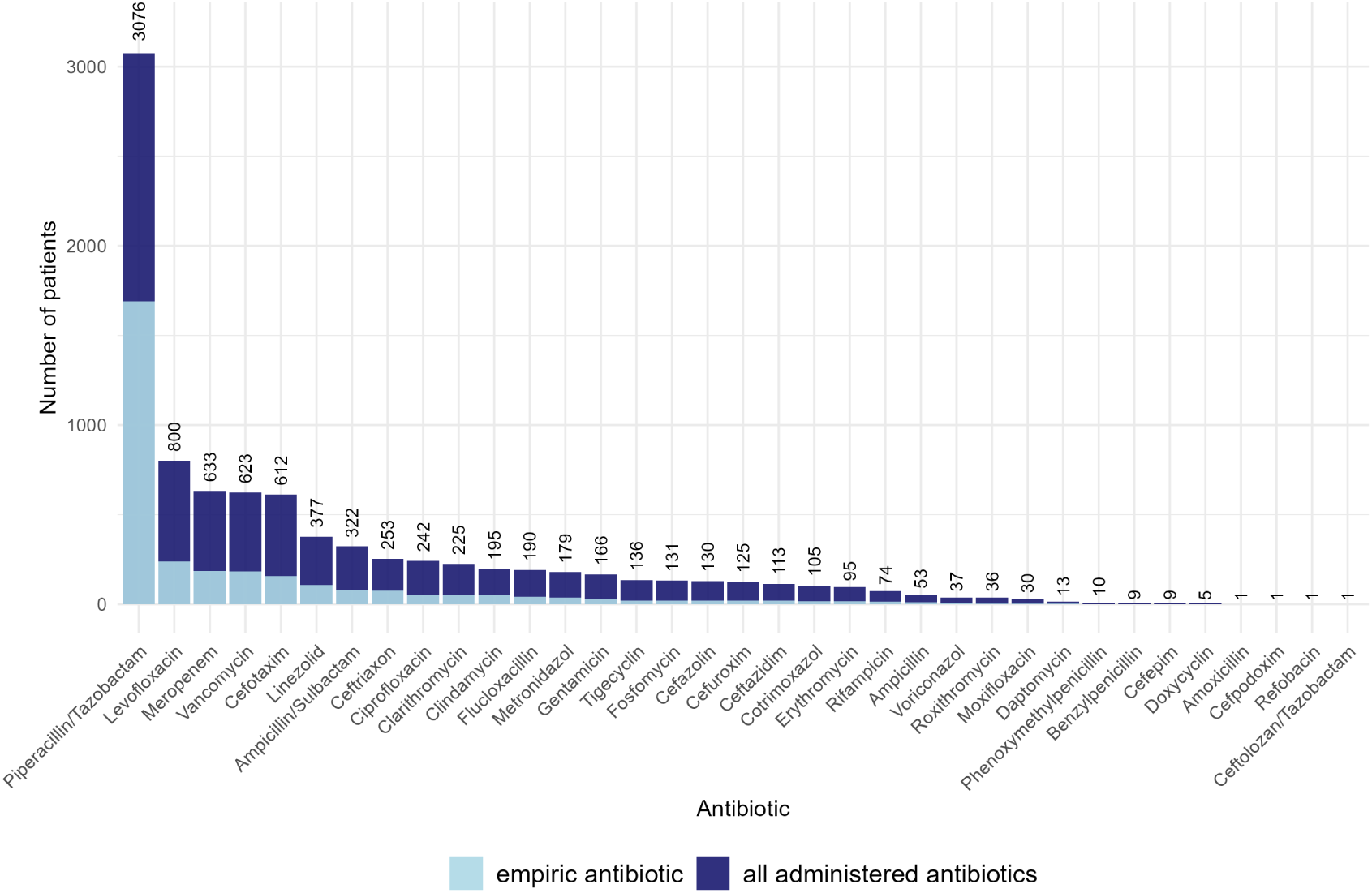
Number of patients who received the respective antibiotic between 2012 and 2023. The light blue bars represent all empiric antibiotic prescriptions (simultaneously administered therapies, i. e. combined therapies, are shown separately), while the dark blue bars additionally include all subsequent administered antibiotics. If antibiotics were administered more than once to one patient, they are counted only once. Printed numbers refer to the dark blue bars.

**Table 1:**
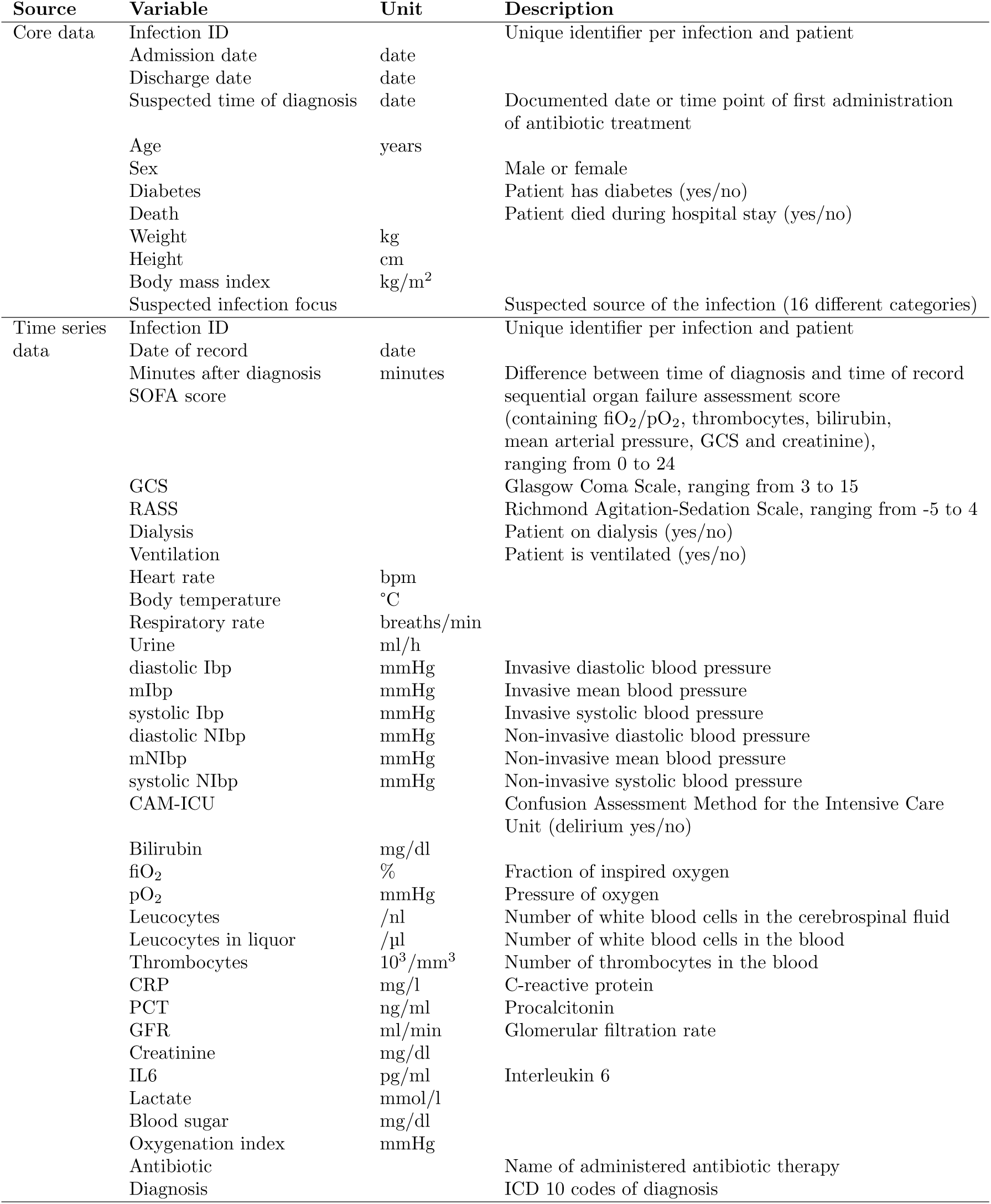
Description of the variables contained in the preprocessed dataset.

A review of the core data reveals that the variables weight, height and body mass index each contain 57 % missing values. In addition, 15 % of patients have no suspected infection focus. However, all other information from the core data is completely available for all patients.

For more effective comparison between patients, we extend the time series data containing laboratory values and vital signs to include the variable ‘minutes after diagnosis’. This is calculated by the difference between the suspected time of diagnosis and the time of record. Importantly, not all variables are constantly measured or measured at equidistant intervals; certain values such as heart rate, however, are frequently measured. Conversely, blood values, including lactate and C-reactive protein (CRP), are typically assessed no more than once per day.

In summary, when considering the average availability of measurements per patient (excluding those with no recorded measurements for a specific value), the values with the lowest frequency of measurement are leucocytes in the liquor, with an average of 0.87 measurements per patient, and procalcitonin (PCT), with an average of 4.10 measurements per patient. In contrast, along with dialysis and ventilation, heart rate (mean of 177.31 measurements per patient) and invasive blood pressure (mean of 160.87 systolic measurements per patient) are the values with highest availability. An overview of the frequency and availability of all time series values is shown in Figure A1 in Appendix A.

### 2.2 Statistical Analysis

Our aim was to develop a CDSS for an empiric antibiotic therapy which is applicable in situations where patients have just been diagnosed with sepsis. Consequently, we intend to predict the prescribed empiric antibiotic therapy based on patients’ core data and health records. We address this goal through statistical classification, where each distinct therapy defines one class, and we train a classifier which assigns class memberships to patients. This is possible since the dataset used for training contains the relevant information about the administered initial antibiotic therapy, i. e. the true class membership.

To start with, we assume a maximum of one measurement per variable for newly diagnosed patients. This mimics the constraints for a rapid decision after sepsis onset. Accordingly, in Section 2.2.1, we reduce our dataset to cross-sectional data and apply a cross-sectional classification model (CCM). In addition, we ask the question how recommendations could have been improved when based on time series data. To that end, we consider time series classification models (TSCM) in Section 2.2.2.

#### 2.2.1 Cross-sectional Classification

Classification is a statistical method for supervised learning, where the true class memberships of training samples are known and can be used during model training to learn a mapping of covariables to classes. After model training, this learned mapping is applied to new data to make predictions for each sample. In our work we rely on the following classifiers for this task: random forests (RF), gradient boosting classifiers (GBC), support vector classifiers (SVC) and multilayer perceptrons (MLP).

To train the CCMs, we further prepare the data additionally to the general preprocessing performed in Section 2.1.2. In particular, we derive covariables which contain cross-sectional instead of time series information as required: for each time-resolved variable, we derive the relevant values for all patients by selecting the first value which has been measured within the first hour after diagnosis. If no measurement is available within this time interval, we use the most recent value within 24 hours before the sepsis diagnosis. Further, we summarize categories of the suspected infection focus occurring less than 50 times to ‘others’.

Many variables in the data have a high proportion of missing values. We refrain from data imputation via, e. g. multiple imputation [39] or filling missing values by the mean across all samples as we want to avoid introducing additional uncertainty into the data. Instead, we discard those variables which have 30 % or more of missing values. Accordingly, we keep the following variables: age, sex, heart rate, average arterial blood pressure, body temperature, fiO_2_, pO_2_, leucocytes, thrombocytes, CRP, creatinine, lactate, blood sugar, bilirubin, requiring dialysis, requiring ventilation and suspected infection focus. These serve as covariables for the CCMs. We do not include systolic and diastolic blood pressure, oxygenation index or glomerular filtration rate as this information is already contained in other variables. After variable selection, we filter for complete cases (i.e., patients without any missing value) and end up with 1194 patients for model estimation. As target variable, i. e. the variable which determines a patient’s class membership, we consider the first administered antibiotic therapy (or a combination of several simultaneously administered therapies) of a patient, i. e. the antibiotic treatment within the first hour after the diagnosis.

In the resulting dataset, we are faced with highly imbalanced class sizes. We mitigate this imbalance by merging antibiotic therapies with less than 20 occurrences to ‘others’. This avoids the CCMs being trained on too few samples for some antibiotics, but even subsequent to this step, imbalanced classes in the target variable remain evident (cf. Table 2): while the largest class, ‘Piperacillin/Tazobactam’, contains 550 patients, the smallest class, namely a combined therapy of ‘Piperacillin/Tazobactam’ and ‘Levofloxacin’, consists of only 21 patients.

**Table 2:** Numbers of antibiotic therapies in the German hospital cohort used for the two-step models.

| **Antibiotic therapy** | **Count** |
| --- | --- |
| Piperacillin/Tazobactam | 550 |
| others | 174 |
| Cefotaxim | 104 |
| Meropenem | 77 |
| Ampicillin/Sulbactam | 75 |
| Levofloxacin | 54 |
| Ceftriaxon | 44 |
| Cefuroxim | 33 |
| Vancomycin | 33 |
| Ciprofloxacin | 29 |
| Piperacillin/Tazobactam - Levofloxacin | 21 |

Common rebalancing techniques like downsampling would lead to a substantial decrease of the sample size. Thus, instead, we divide the original classification task into two consecutive tasks: in a first step, we perform binary classification to predict the majority class ‘Piperacillin/Tazobactam’ versus all others; in a second step, we perform multiclass classification to further distinguish between the remaining antibiotic therapies. As described above, we apply RF, GBC, SVC, and MLP as classifiers at both steps.

To account for the fairly small amount of data and to avoid model overfitting, we use nested cross-validation (CV) with an inner and an outer loop [40]. The outer loop consists of ten folds and splits the whole dataset into ten disjunct sets. The inner loop consists of five folds allowing for a grid search to optimize the models’ hyperparameters. For RF, we tune the splitting criterion, the total number of fitted trees, the maximum depth of the trees and the minimum number of samples to perform a subsequent split. For GBC, in addition to the latter two parameters, we optimize the learning rate as well as the number of boosting stages. For SVC, the regularization parameter *C*, the kernel type and the degree in case of a polynomial kernel are tuned during the grid search. For MLP, we optimize the number of hidden layers assuming each layer to consist of 100 neurons, the activation function, the solver, the *L*2 regularization term and the initial learning rate. To measure the models’ performance, we use the F1-score, which is the harmonic mean between precision and sensitivity.

Estimating the binary model does not require any rebalancing. The dataset (before applying CV) contains 550 patients receiving ‘Piperacillin/Tazobactam’ (cf. Table 2) and henceforth 644 patients receiving a different therapy. This is, however, different for the multiclass classification in the second step. The model is trained on those patients from the training set who received an antibiotic therapy other than ‘Piperacillin/Tazobactam’. As shown in Table 2, there are still large differences between the remaining ten class sizes. At this point, after pre-empting the most dominant class ‘Piperacillin/Tazobactam’, we opt for rebalancing to prevent the model from ignoring the minority classes. As the difference between the largest and smallest class size is substantial, we downsample the classes the sizes of which exceed the average frequency in the training data for the second step and upsample the other classes accordingly. This procedure prevents the dataset from becoming too small, but at the same time we avoid obtaining too many replicates.

#### 2.2.2 Time Series Classification

We apply a TSCM to investigate whether more information is beneficial to correctly predict an empiric antibiotic therapy for sepsis patients. In contrast to the models described above, TSCMs consider a time series of a sample instead of using just one measurement per variable. In the medical context, this poses a particular challenge, given that patients’ health records are typically measured at different time points rather than at uniform intervals. Consequently, the individual time series often vary in length. This requires techniques which can deal with unequal sequences, such as the *k*-nearest neighbor (*k*-NN) algorithm employing dynamic time warping (DTW) to measure distances.

To apply *k*-NN with DTW to our data, we consider the patients’ time series starting at 24 hours before sepsis diagnosis and up to one hour afterwards. Our approach is based on the fact that no later measurements are available for new patients due to the need for fast antibiotic treatment. We use the same covariables and target variable as for the CCMs described above. In contrast to these models, we cannot exclusively use complete cases for the TSCM, as the data basis would become far too small. Therefore, we use forward or, if no previous value is available, backward propagation to fill the missing values in the time series. The procedure to estimate the models is the same as for the CCMs: in a first step, we perform binary classification to derive a probability for ‘Piperacillin/Tazobactam’, and in a second step, we aim at classifying those antibiotic therapies that are different from ‘Piperacillin/Tazobactam’. Again, we use nested CV and a combined procedure of up- and downsampling for class rebalancing in the second step.

## 3 Results

### 3.1 Requirements and Insights from Clinical Knowledge

To enhance the reliability of the CDSS, it was important for us to define required and potentially influential information prior to the data analysis, and to acknowledge limitations. In general, there is a wide variety of variables in our dataset, and in a purely data-driven approach, we could have allowed the system to search for patterns in an unrestricted manner. However, this carries the risk of spurious correlations or therapies that may turn out impractical. Through the interviews and discussions described in Section 2.1.1, the following facts and requirements were compiled and subsequently served as a knowledge base for the non-clinical scientists in this project.

### Including relevant factors for treatment decision

Following a diagnosis of sepsis, there are multiple factors that the treating physician takes into consideration when selecting an appropriate antibiotic. These include, among others, core data, measured vital signs and blood values, patient allergies, known co-morbidities, adverse effects and image data (e. g. X-ray images or computer tomography scans). Further, the identification of the infection focus is of utmost importance as it allows for the identification of potential pathogens once the affected organ is confirmed. However, in some cases the focus is not provided as it is not directly recognizable.

### Considering risk factors for antibiotic resistances

In order to take potential resistances into account, whether on a global or patient-specific basis, it is necessary to consider additional kinds of information: the patient’s origin and professional background as well as their travel history, previous hospitalization or stay in nursing homes, and antibiotic intake within the last three to six months. This information may offer insights into the present pathogens and suspected resistances for specific patients.

### Evaluation of antibiotic effectiveness

With regard to the evaluation of antibiotic effectiveness, i. e. the assessment of whether the administration of a specific antibiotic was successful in treating a particular case of sepsis, the most important information are the SOFA score, oxygenation index, PCT, CRP, interleukin 6 (IL6), body temperature and circulatory parameters such as blood pressure, heart rate, and respiratory rate. From these, especially PCT and IL6 are also identified in the literature [41]. Furthermore, leucocytes, thrombocytes, bilirubin, creatinine, lactate, CAM-ICU and urine output are considered as potential covariables. Generally, for an effective therapy, laboratory values should show a clear improvement. However, the clinical experts emphasized that the identification of relevant factors depends on the infected organ. Furthermore, they highlighted that the general health condition of a patient is more important than laboratory values. If a patient’s well-being improves, irrespective of laboratory values, and they demonstrate, e. g. increased responsiveness and alertness, it can be assumed that the administered therapy was effective.

### Expectations and acceptance of CDSS

One of the conducted interview-series addressed the physicians’ opinion on the practical implementation of a CDSS and revealed high interest and need for the support by such a system. This is because clinicians must process the available information within a short time frame, often facing incomplete data when the critical health status of a patient necessitates a rapid therapeutic decision. Furthermore, increasing antimicrobial resistances demand careful consideration of the numerous alternative therapies available. The establishment of a support system, therefore, is of great added value for physicians with limited practical experience, thereby helping them to make well-informed treatment decisions. However, it was also noted that there are still many operational hurdles to overcome, such as the lack of digitization in clinics or the supply of reliable and reasonable retrospective data, which hinders its use in practical hospital settings. A detailed analysis of the interviews is provided by [14]. Another important requirement for a CDSS highlighted by the medical professionals concerned the interpretability of the underlying statistical models. Since we employ interpretable classification methods rather than AI tools here, this constraint is met but must be given careful consideration in general.

### 3.2 Challenges from Data Availability

A goal of our study was to determine the extent to which the ideal and theoretically formulated requirements for a CDSS align with the reality of clinical practice. The result of our investigation was that we encountered a number of challenges regarding data quality and availability, which partially led to questions not being answered to the extent we had originally envisioned. These challenges are not issues that exist solely at our site but arise generally from the common level of data documentation in everyday clinical practice and the prospective collection of data for research purposes, influenced by the degree of digitization of clinical data in German hospitals. We provide a detailed account of the most impactful difficulties here to highlight the necessities, and we describe how we overcame some of them by identifying alternative solutions.

One major limitation was the amount of data. While we had originally planned to export data from all three consortium hospitals, this was made impossible due to technical limitations as well as differing processes across the hospitals. As a consequence, the present study is constrained to data of one single hospital. This resulted in the rather small sample size described before, and this in turn excluded AI-based models which require much larger amounts of data [42].

A further substantial restriction resulted from the fact that several variables, which had been identified as highly important, were not present in the dataset. These included patient history information from which one could have derived the risk of individual resistances against certain antibiotics, such as origin, profession, travel history and previous antibiotic prescriptions. Moreover, we lacked information on the occurrence of adverse effects, which was not even provided for severe cases. Consequently, our investigation could not address the identification of antibiotic therapies with minimal adverse effects.

Furthermore, the suspected infection focus, which was highlighted by the doctors as one of the most relevant factors in the decision-making process for an antibiotic therapy, was not routinely documented. In order to still obtain a realistic prediction, patient diagnoses associated with the infection foci were used to infer the suspected infection focus for individual patients (cf. the preprocessing steps outlined in Section 2.1.2).

Next, the exact time of diagnosis was unknown in the sense that its temporal alignment was partially shifted when the documentation started after the first treatment. Hence, there is uncertainty regarding the patient’s condition at the time of diagnosis.

Regarding the effectiveness of an antibiotic therapy, the medical experts asserted that the clinical impression of a patient was given primacy over laboratory values. Still, no such data was documented. Thus, no direct quantitative measure was available to validate the effectiveness of the given therapies.

Even for those variables that were theoretically included in the dataset, there were many missing values. Thus, despite the ongoing digitization in hospitals and the consideration of retrospective data from more than ten years, the final sample size of complete patient data was relatively small as reported in Section 2.1.2.

Another challenge, particularly in the context of time series analyses, pertains to the fact that the longitudinal measurements are obtained at varying time intervals per patient and with differing frequencies among the measured values. The generation of the variable ‘minutes after diagnosis’ (cf. Section 2.1.2) resulted in a data matrix for longitudinal values that was extremely sparse. This issue was addressed as described in Section 2.2.

Concerning the primary target variable, the empiric antibiotic therapy, we encountered high class imbalance (cf. Figure 2). The ten most frequently prescribed antibiotics accounted for more than 89 % of all empiric antibiotics, with ‘Piperacillin/Tazobactam’ being the most frequently administered therapy, accounting for 54 % of all prescriptions. This imbalance in prescriptions posed a major challenge in predicting outcomes for underrepresented classes and may have lead to biased classification results [43]. We addressed this issue with a two-step modeling approach as outlined in Section 2.2.

To evaluate our classification approach beyond the previously described dataset and to gain insights into the generalizability of our method, we sought to incorporate an additional dataset. The most suitable source for this purpose was the MIMIC IV database (Medical Information Mart for Intensive Care, [44]), which contained health-related data from patients admitted to the ICU of the Beth Israel Deaconess Medical Center (Boston, Massachusetts) between 2008 and 2019. However, important information such as the infection focus or PCT measurements were missing in the database. Further, we detected notable discrepancies between Germany and the USA with regard to the fundamental hospital procedures and guidelines for the management of sepsis, as well as the related data basis (e. g. [45]), which disqualified the MIMIC IV database as a solid basis for comparison. Thus, in order to develop a targeted approach for common practice in Germany, we exclusively relied our analysis on the data described in Section 2.1.2 and emphasize the need for more and better data about the diagnosis and treatment of sepsis patients.

### 3.3 Learnings from Statistical Modeling

#### 3.3.1 Cross-sectional Classification

Despite the aforementioned challenges, we used various approaches to statistically model the physicians’ decision for an empiric antibiotic therapy. Tables 3 and 4 show the results of the cross-sectional classification models (RF, GBC, SVC, MLP; cf. Section 2.2.1). The results from both tables refer to the same model prediction but are split into two parts for better readability (each table displays five to six therapies). Due to the outer loop of the nested CV, we obtain ten predictions for different test sets; the tables present their averages.

**Table 3:**
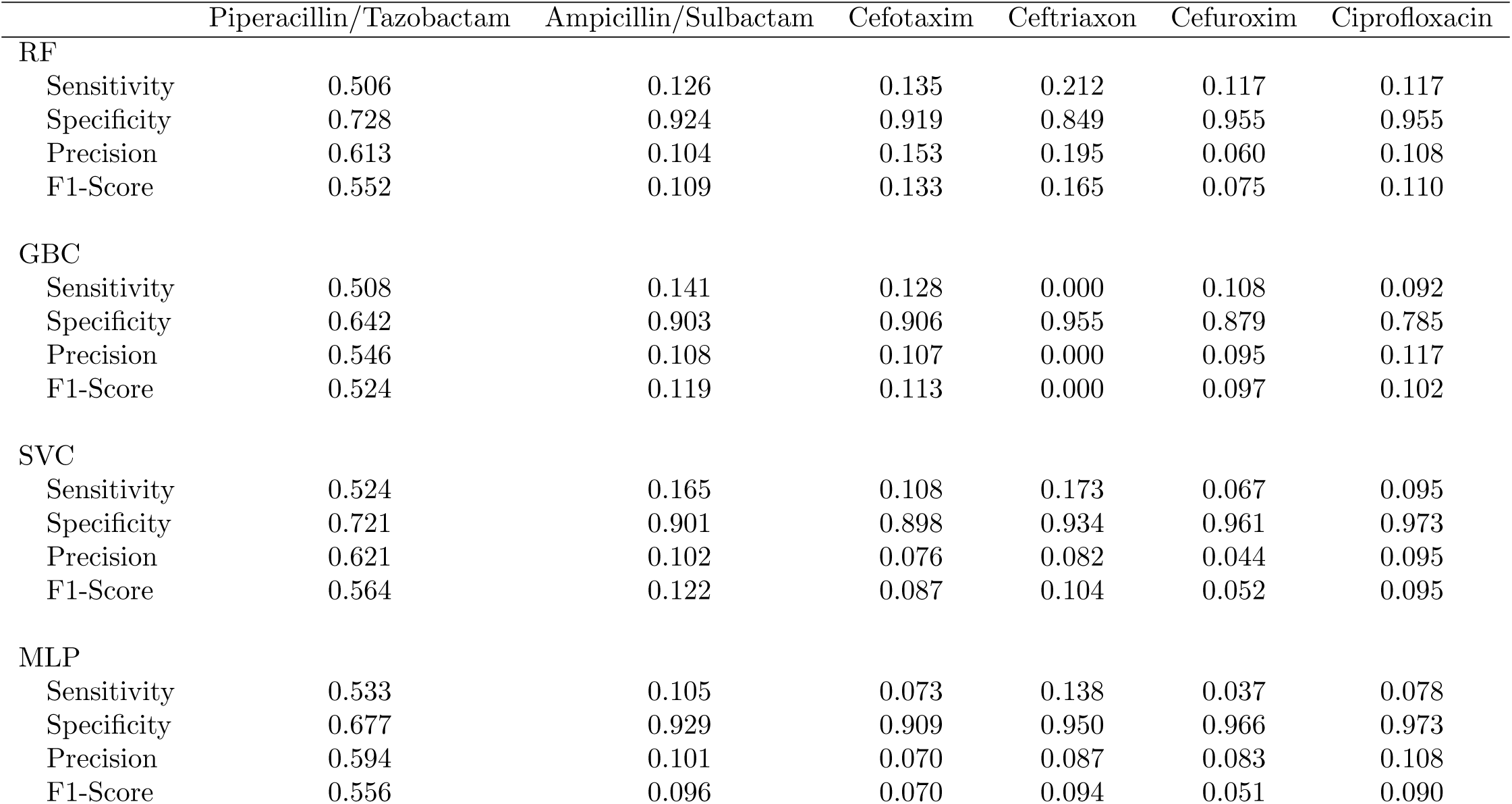
Average performance of the CCMs across ten folds of nested CV for considered antibiotics (part 1).

**Table 4:** Average performance of the CCMs across ten folds of nested CV for considered antibiotics (part 2).

|  | Levofloxacin | Meropenem | Piperacillin/Tazobactam-Levofloxacin | Vancomycin | others |
| --- | --- | --- | --- | --- | --- |
| **RF** |  |  |  |  |  |
| Sensitivity | 0.184 | 0.116 | 0.000 | 0.125 | 0.116 |
| Specificity | 0.918 | 0.912 | 0.978 | 0.962 | 0.945 |
| Precision | 0.112 | 0.099 | 0.000 | 0.051 | 0.265 |
| F1-Score | 0.135 | 0.104 | 0.000 | 0.069 | 0.153 |
| **GBC** |  |  |  |  |  |
| Sensitivity | 0.083 | 0.143 | 0.033 | 0.050 | 0.118 |
| Specificity | 0.939 | 0.907 | 0.586 | 0.869 | 0.919 |
| Precision | 0.055 | 0.099 | 0.033 | 0.050 | 0.198 |
| F1-Score | 0.061 | 0.112 | 0.033 | 0.050 | 0.142 |
| **SVC** |  |  |  |  |  |
| Sensitivity | 0.042 | 0.107 | 0.083 | 0.120 | 0.112 |
| Specificity | 0.933 | 0.934 | 0.880 | 0.964 | 0.937 |
| Precision | 0.033 | 0.109 | 0.045 | 0.064 | 0.249 |
| F1-Score | 0.037 | 0.104 | 0.058 | 0.072 | 0.152 |
| **MLP** |  |  |  |  |  |
| Sensitivity | 0.081 | 0.156 | 0.000 | 0.145 | 0.157 |
| Specificity | 0.937 | 0.934 | 0.874 | 0.949 | 0.925 |
| Precision | 0.064 | 0.122 | 0.000 | 0.052 | 0.268 |
| F1-Score | 0.070 | 0.132 | 0.000 | 0.069 | 0.191 |

It becomes immediately apparent that none of the four models achieved results that meet the requirements we expect for a reliable model: for most antibiotics, the sensitivity is lower than 22 %, in many cases it is even lower than 10 %. This low sensitivity comes along with a high specificity. The only exception is ‘Piperacillin/Tazobactam’, for which all four classification models result in a sensitivity above 50 % and a specificity above 60 %. MLP achieved the highest sensitivity of 53.3 %; this is accompanied by a medium-high specificity of 67.7 %. Compared to all other antibiotic therapies, the performance of the four models for predicting ‘Piperacillin/Tazobactam’ are satisfactory: both sensitivity and specificity, and thus precision and F1-score, are moderately high. However, this performance is still insufficient for clinical implementation.

#### 3.3.2 Time Series Classification

The averaged results from time series classification across the ten folds of the CV are shown in Table 5. No improvement of CCM results could be achieved using time series data. *k*-NN with DTW performed worse than the CCMs as measured by F1-score. Even for ‘Piperacillin/Tazobactam’, the F1-score was on average 12.3 percentage points lower when using the TSCM instead of CCMs. The poor performance for ‘Ciprofloxacin’ and ‘Vancomycin’ is conspicuous. In case of ‘Ciprofloxacin’, the sensitivity is low, and so is the specificity. For ‘Vancomycin’, not a single sample belonging to this class was correctly classified.

**Table 5:**
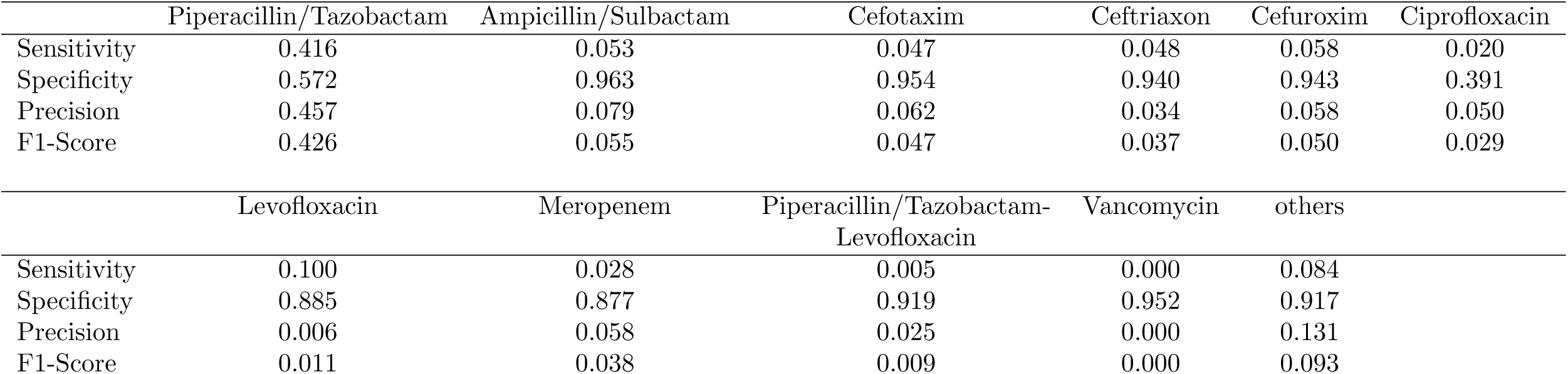
Average performance of the TSCM across ten folds of nested CV for all antibiotics.

|  | Piperacillin/Tazobactam | Ampicillin/Sulbactam | Cefotaxim | Ceftriaxon | Cefuroxim | Ciprofloxacin |
| --- | --- | --- | --- | --- | --- | --- |
| Sensitivity | 0.416 | 0.053 | 0.047 | 0.048 | 0.058 | 0.020 |
| Specificity | 0.572 | 0.963 | 0.954 | 0.940 | 0.943 | 0.391 |
| Precision | 0.457 | 0.079 | 0.062 | 0.034 | 0.058 | 0.050 |
| F1-Score | 0.426 | 0.055 | 0.047 | 0.037 | 0.050 | 0.029 |

|  | Levofloxacin | Meropenem | Piperacillin/Tazobactam-Levofloxacin | Vancomycin | others |
| --- | --- | --- | --- | --- | --- |
| Sensitivity | 0.100 | 0.028 | 0.005 | 0.000 | 0.084 |
| Specificity | 0.885 | 0.877 | 0.919 | 0.952 | 0.917 |
| Precision | 0.006 | 0.058 | 0.025 | 0.000 | 0.131 |
| F1-Score | 0.011 | 0.038 | 0.009 | 0.000 | 0.093 |

## 4 Discussion

### 4.1 Exploration of Model Performance

Despite careful data preparation, modeling, and incorporation of clinical knowledge, the statistical models achieved only poor to moderate performance in predicting the physicians’ decision for an empiric antibiotic therapy. Only the first step of the two-step approach, i. e. the classification of ‘Piperacillin/Tazobactam’ against all other therapies, succeeded to some extent. We identify several reasons that we suspect to be contributing to this issue.

### Sample size

Good predictions require sufficient sample sizes. Our final dataset contained only 1194 samples in total. 550 out of these belonged to the class ‘Piperacillin/Tazobactam’; all other class sizes ranged from 21 to 174. The comparably good performance in the first step of the classification procedure indicates that, all in all, our statistical models are appropriate as long as enough data is available. To put it another way, we suspect the main reason for the poor overall predictive ability being the small sample and class sizes. Sepsis manifests itself in very different ways; to find similarities in patients’ health records, many more samples may be needed to be considered.

### Broad-spectrum antibiotics

Notably, the most commonly prescribed therapy, ‘Piperacillin/Tazobactam’ (roughly 50 %), along with several other documented treatments, are broad-spectrum antibiotics. This reflects how often this kind of antibiotics is prescribed in clinical practice. In some cases, this antibiotic therapy is recommended by the guidelines; however, the high frequency of prescriptions also suggests a paucity of relevant information for a more targeted treatment decision.

### Class imbalance

Not only was the sample size small, but the classes were also substantially different in size. The over-representation of ‘Piperacillin/Tazobactam’ led to a notable class imbalance. To deal with this, we decided to split the classification task into two steps. For the first step, the prediction of ‘Piperacillin/Tazobactam’, no class rebalancing was needed, and we achieved comparably good results using CCMs. Still, these results are not yet suitable for clinical practice, highlighting the need for large high-quality datasets, in particular for AI-based classifiers. In the second step, in contrast, we had to rebalance the classes as these were still unequally represented. Depending on the method used for rebalancing, useful information was removed (e. g. when using random undersampling of the majority class [46]), samples were duplicated (e. g. when using random oversampling of the minority class [46]) or synthetic data was added, potentially containing unrealistic or even wrong information (e. g. when using Synthetic Minority Over-sampling Technique [47]). Consequently, the application of methods for class rebalancing comes with uncertainty. This, in conjunction with the small sample size, is a possible explanation for the poor predictive ability of the models.

### Observed initial treatments

In order to model a physician’s decision for an empiric antibiotic therapy, we used the first administered antibiotic contained in the dataset. This happened irrespectively of whether the therapy was effective or not because the dataset does not contain quantified information about the effectiveness (cf. Section 3.2). However, there are indications that the first antibiotic was not necessarily the most effective one and could thus lead to inappropriate predictions: the data shows that, due to constant re-evaluation, the administered therapy was modified several times during one infection for many patients. If appropriate information became available about the effectiveness, the target variable could, however, be easily adapted.

### Missing information

As outlined in Section 3.2, relevant variables for an informed decision in favor of an antibiotic therapy were not included in our data. In particular, there were no variables that provide information needed to assess a patient’s risk for suffering from resistant bacteria, such as the origin, the travel history and the profession. Knowledge about such possible risk factors is crucial to limit the eligible antibiotic therapies. Without this information, the model may not be able to predict an antibiotic therapy which is consistent with the empiric administered therapy chosen by the physician.

### Uncertain infection focus and time of diagnosis

Medical experts regarded the infection focus to be of high importance within the decision-making process for an antibiotic therapy (cf. Section 3.1). In Section 2.1.2, we explained how we derived the suspected focus. This procedure, however, comes with uncertainty, and we consider possible errors to further affect the models’ performance. A similar statement holds for the time point of sepsis diagnosis as derived by us.

### Missing data

Many classifiers are unable to deal with missing values. Thus, we reduced the dataset to complete cases by removing certain covariables and patients. This further limits the classification models’ interpretability and increases prediction uncertainty.

In light of all the aforementioned limitations, the poor classification performance especially in the second step of our approach becomes comprehensible. The shortcomings highlight the need for large high-quality data as a basis for a reliable CDSS for the individualized prediction of a targeted effective antibiotic therapy for patients diagnosed with sepsis.

### 4.2 Conclusion

We embarked on an ambitious project to develop a CDSS aimed at recommending an appropriate initial antibiotic therapy for patients with sepsis. This CDSS was intended to be applicable immediately upon diagnosis and to provide recommendations regarding potential resistances and side effects. The system was to be developed using data science methods, drawing from data collected at three local hospitals.

However, the situation within the clinics revealed that various factors contributed to a relatively small dataset, comprising only 1194 complete cases, along with a substantial amount of unavailable information. Contemporary AI approaches typically require large datasets, therefore, a swift implementation using large language models, which may seem straightforward to an outsider, was not feasible in this context. Instead, we took on the challenge by combining meticulous information and data preparation with the adaptation of statistical learning methods. In fact, we achieved a satisfactory classification for one of the sub-questions. Nonetheless, our project uncovered several limitations, and in this article, we outline our key insights regarding the process.

To summarize, the process included the initial stages of knowledge acquisition, data preprocessing, overcoming data-related challenges, and constructing predictive models to classify the physician’s choice of empiric therapies. We have described each relevant step, examined the challenges involved, and presented potential models together with the results derived from the available data. Our work yields two principal take-home messages: first, the importance of interprofessional collaboration, and second, the need for high-quality data and improvements in data acquisition processes.

Regarding the first, the interprofessional collaboration and the continuous exchange between medical experts and model constructors turned out to be crucial both on preliminary stages of the project and during its entire progress. Such collaboration increases the chance of obtaining meaningful results as all stakeholders are aware of the necessary requirements for data availability and quality. Many medical relationships require detailed discussion before they can be integrated into the data and modeling processes. Simultaneously, critical questions — often stemming from one party’s lack of expertise — can lead to new insights and perspectives.

Regarding the second, the results of the models, discussed in Section 4, highlight the critical need for comprehensive and large high-quality data. Our dataset contains information on patients admitted to the ICU, where the patients are constantly monitored. Here, certain parameters are measured in an automated way, while the measurement of many other values has to be actively initiated and is carried out manually. The latter may lead to a lack of continuity in health records, and it is expected that data from less controlled units (like non-ICU) is of even lower quality. To improve the general documentation of health data, the optimization and expansion of digitization in German hospitals is of highest importance. The implementation of the electronic patient record system (ePA, [48]), which is currently being introduced in Germany, has the potential to offer great benefit to scientific analyses. Specifically, information regarding patients’ origin, travel history, professional background, and time of diagnosis could be made more readily accessible. Even information from previous inpatient or outpatient stays can potentially provide relevant information that would be available through the ePA. The benefit of the ePA for research purposes is only provided if access to the data is granted. The draft of the German ‘Forschungsdatengesetz’ [49] aims at improving access grant to and use of data for research purposes; consequently contributing to an improvement in the data availability. Moreover, a health data usage act (‘Gesundheitsdatennutzungsgesetz’, [50]) was passed in March 2024 in Germany, with the objective of regulating the use of health data for research purposes in the public interest and for the data-based further development of the healthcare system.

The logical next steps that arise from our project are to move beyond mere classification of therapies that were prescribed in practice to providing recommendations for therapies that are expected to have high effectiveness. To overcome the lack of a quantitative measure for administered antibiotic therapies, this may involve the data-driven assessment of the health status of sepsis patients from time series of laboratory values and vital signs. This may provide a basis for the development of methods that can learn from retrospective data to predict the best possible therapy outcome for an individual sepsis patient. The steps achieved and presented here will contribute to the development of an effective CDSS for antibiotic prescription in sepsis, ultimately improving the survival of sepsis patients and helping to reduce antibiotic resistance.

### 4.3 Computational Details

The data preprocessing was done using the statistical software R version 4.4.2 [51]; the models were fitted using the statistical software Python version 3.11.9 [52]. The code used for model fitting is available at https://github.com/fuchslab/CDSS_Sepsis.

## Data Availability

Due to data privacy protection, we are not allowed to share the data of the German hospital used in this work.

https://github.com/fuchslab/CDSS_Sepsis

## Acknowledgments

We thank all members of the ‘KINBIOTICS’ project, especially the medical experts who took part in the interviews and meetings and thus made an important contribution to the model development. This study was supported by a grant from the German Federal Ministry of Health (Bundesministerium für Gesundheit), grant ZMVI1-2520DAT930 (Kinbiotics).

## Abbreviations

AI: artificial intelligence
ANN: artificial neural network
CCM: cross-sectional classification model
CDSS: clinical decision support system
CIS: clinical information system
CRP: C-reactive protein
CV: cross-validation
DT: decision tree
DTW: dynamic time warping
EvKB: Evangelisches Klinikum Bethel
GBC: gradient boosting classifier
ICU: intensive care unit
IL6: interleukin 6
MLP: multilayer perceptron
PCT: procalcitonin
RF: random forest
SOFA: sequential organ failure assessment
SVC: support vector classifier
TSCM: time series classification model
WHO: World Health Organization

## Appendix A Missing Values

**Figure A1:**
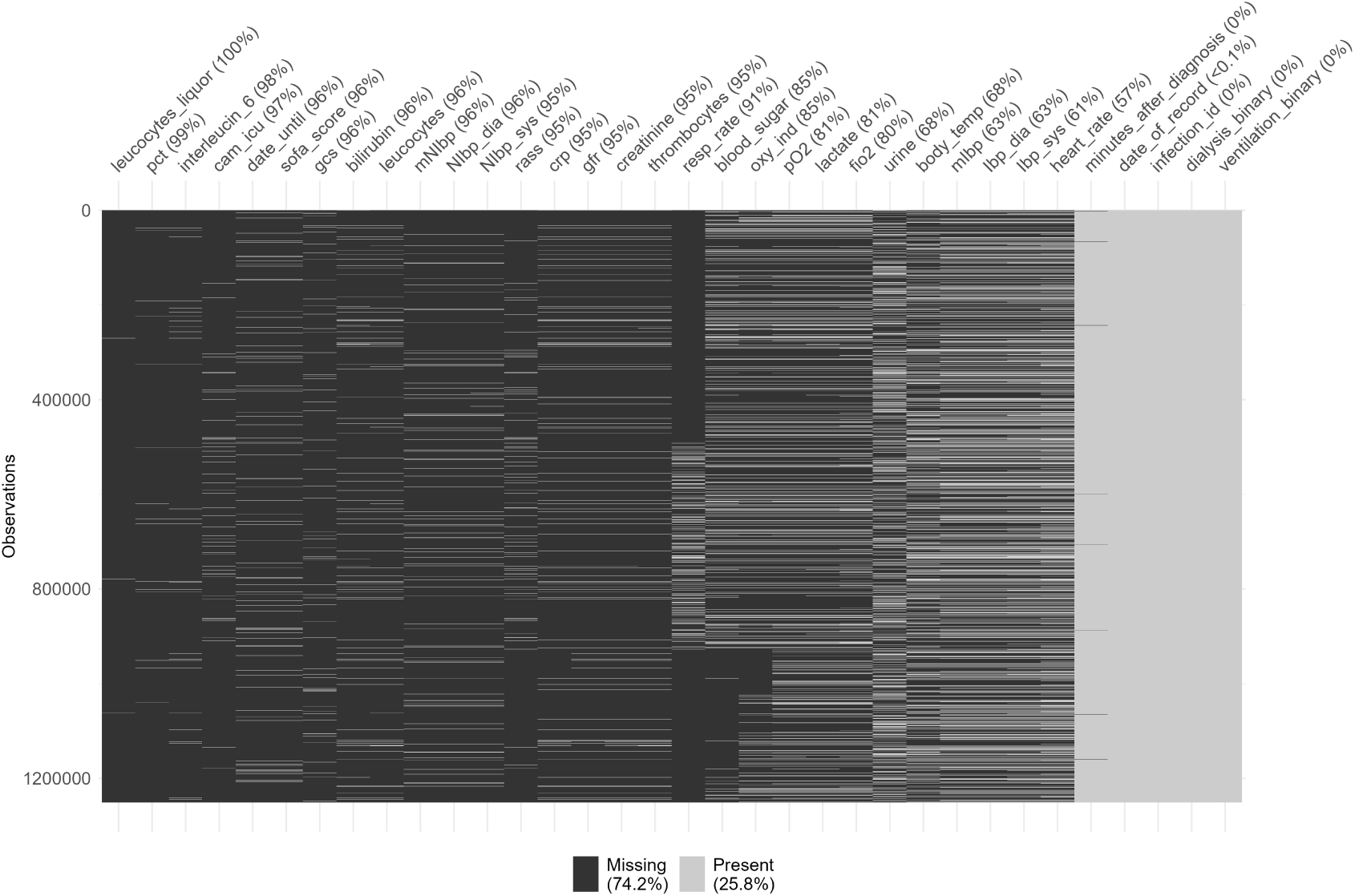
Percentages and pattern of missing values in time series variables. Observations are shown in rows, variables in columns. Black areas are missing values while gray areas reflect available entries.

